# Quantification of blood glial fibrillary acidic protein using a second-generation microfluidic assay. Validation and comparative analysis with two established assays

**DOI:** 10.1101/2023.08.24.23294528

**Authors:** Badrieh Fazeli, Nerea Gómez de San José, Sarah Jesse, Makbule Senel, Patrick Oeckl, Deborah K Erhart, Markus Otto, Steffen Halbgebauer, Hayrettin Tumani

## Abstract

**Background:** Increased levels of glial fibrillary acidic protein (GFAP) in blood have been identified as a valuable biomarker for some neurological disorders, such as Alzheimer’s disease and multiple sclerosis. However, most blood GFAP quantifications so far were performed using the same bead-based assay, and to date a routine clinical application is lacking.

**Methods:** In this study, we validated a novel second-generation (2^nd^ gen) Ella assay to quantify serum GFAP. Furthermore, we compared its performance with a bead-based single molecule array (Simoa) and a homemade blood GFAP assay in a clinical cohort of neurological diseases, including 210 patients.

**Results:** Validation experiments resulted in an intra-assay variation of 10%, an inter-assay of 12%, a limit of detection of 0.9 pg/mL, a lower limit of quantification of 2.8_pg/mL, and less than 20% variation in serum samples exposed to up to five freeze-thaw cycles, 120_hours at 4 °C and room temperature. Measurement of the clinical cohort using all assays revealed the same pattern of GFAP distribution in the different diagnostic groups. Moreover, we observed a strong correlation between the 2^nd^ gen Ella and Simoa (r=0.91 (95% CI: 0.88 - 0.93), p<0.0001) and the homemade immunoassay (r=0.77 (95% CI: 0.70 - 0.82), p<0.0001).

**Conclusions:** Our results demonstrate a high reliability, precision and reproducibility of the 2^nd^ gen Ella assay. Although a higher assay sensitivity for Simoa was observed, the new microfluidic assay might have the potential to be used for GFAP analysis in daily clinical workups due to its robustness and ease of use.

**What is already known on this topic:**

- Blood glial fibrillary acidic protein (GFAP) levels are an emerging biomarker for diagnosing, prognosis and treatment monitoring for AD, MS and other neurological disorders. However, so far, the application in clinical routine remains a challenge.

**What this study adds:**

- This study validated a novel, easy-to-use second-generation microfluidic assay for the quantitative measurement of blood GFAP. Moreover, its performance was compared to two other GFAP immunoassays, including single molecule array.

**How this study might affect research, practice or policy:**

- This study proved the reliability, precision and reproducibility of the novel second-generation microfluidic assay, which might be more easily implemented in daily clinical routine analyses and therefore facilitates the application of GFAP as a biomarker for neurological diseases.

## Introduction

Human glial fibrillary acidic protein (GFAP) is a 432-amino acid long polypeptide encoded by the corresponding gene on chromosome 17q21 (1). It belongs to the type-III intermediate filaments and is responsible for maintaining the mechanical strength of astrocytes which support and regulate the blood-brain barrier (2). Moreover, GFAP is involved in fundamental and critical astrocytic functions like motility, proliferation, synaptic plasticity, myelination and responses to brain damage (3). GFAP is highly but not exclusively expressed in astrocytes in the central nervous system (4).

Reactive astrogliosis is considered to be a consequence of neurodegeneration and neuronal death and refers to morphological and functional changes in astrocytes followed by proliferation and up-regulation of GFAP (5). Because GFAP concentrations are higher in cerebrospinal fluid (CSF) than in blood, any changes in GFAP concentration are easier to detect in CSF. However, several studies demonstrated a higher discriminative power for blood compared to CSF GFAP for several diagnostic groups compared to control patients (6–8).

GFAP has recently drawn attention due to its potential as a promising biomarker for several neurological disorders where it has been shown to have value in disease diagnosis as well as disease progression and treatment monitoring (6,9). Several studies have demonstrated elevated GFAP levels in Alzheimer’s disease (AD)(8,10,11). A recent meta-analysis compared AD patients to healthy controls as well as Aβ-positive to Aβ-negative groups. The findings display a significant increase in blood GFAP levels confirming the diagnostic value of GFAP in AD (12). Furthermore, blood GFAP levels seem to increase more than 10 years before symptom onset in genetic AD patients (13,14). In addition, blood GFAP levels might be able to predict the conversion from AD mild cognitive impairment to AD dementia (10). In multiple sclerosis (MS) GFAP levels vary by MS subtype and may be used as disease severity and progression biomarker (6,15,16).

Recent advancements in highly sensitive technologies facilitated the evaluation of GFAP in several neurological conditions. However, a substantial proportion of these investigations relied upon the same single-molecule array (Simoa) platform (17–19). Therefore, these data need to be validated with an independent assay for potential use in daily clinical routine. Furthermore, efforts to enhance the ease of use of existing techniques remain essential.

In this study, we validated the performance of the novel 2^nd^ gen commercial Ella assay for the assessment of serum GFAP. Furthermore, serum samples of a clinical cohort of 210 patients were selected to measure their GFAP levels using the 2^nd^ gen Ella assay, the Simoa GFAP discovery kit and a sensitive homemade Ella GFAP blood assay (20). The data enabled a comparative analysis of the three assays in terms of GFAP levels in the diagnostic groups, their correlation and agreement.

## Methods

### Patients’ samples

In this study, 210 serum samples from seven diagnostic groups were analyzed. The samples were collected in the Department of Neurology of Ulm University Hospital between 2010 and 2021. All patients or their legal proxies were informed and signed the consent for inclusion in this study. The study was approved by the local Ethics committee from the University of Ulm (approval number: 20/10) and conducted following the Declaration of Helsinki. The clinical cohort included AD (n=44), MS (n=38), behavioral variant frontotemporal dementia (bvFTD) (n=14), Encephalitis (n=6), Meningitis (n=9), Meningoencephalitis (n=4) and control patients (Con) (n=95). AD and MS patients were diagnosed according to the International Working Group 2 criteria (21) and the 2017 revision of the McDonald criteria (22), respectively. For bvFTD diagnosis the international criteria were used (23,24). Encephalitis was diagnosed using the criteria of the International Encephalitis Consortium (25). Viral/unknown origin meningitis patients were identified by taking into account clinical symptoms of meningitis as well as CSF analysis (pleocytosis ≥ 5/µl, blood CSF barrier dysfunction, elevated lactate, possible intrathecal IgG/IgM/IgA synthesis or oligoclonal bands in CSF only). Virus detection was performed by PCR or by testing antibodies in CSF and serum. In cases of meningitis of unknown etiology, pathogen detection was not possible.

The MS cohort consists of patients with clinically isolated syndrome (CIS n=4), relapsing-remitting MS (RRMS n=30), secondary progressive MS (SPMS n=2), and primary progressive MS (PPMS n=2). 95 Con patients were admitted to the hospital with tension-type headaches, temporary sensory symptoms and dizziness. However, neurodegenerative or neuroinflammatory conditions were ruled out after clinical and radiological evaluation. All Con patients underwent a lumbar puncture to exclude an acute or chronic inflammation of the central nervous system (CNS). This evaluation encompassed criteria such as normal leukocyte count, intact blood-CSF barrier function (i.e., normal Albumin CSF-serum ratio), and absence of intrathecal immunoglobulin synthesis (incl. quantitative analysis of IgG, IgA, IgM, and oligoclonal IgG bands).

### Sample collection and analysis

To collect serum samples, venous blood was centrifuged at 2000 g for 10 min and stored within 30 min at -80°C. For stability testing, CSF was also analyzed. For this purpose, CSF samples were centrifuged at 2000 g for 10 min and aliquots were stored within 30 min at -80°C. GFAP levels were then analyzed using the 2^nd^ gen Ella assay, Simoa assay, and a homemade Ella assay. Disease groups were randomized during measurements and two serum quality control (QC) samples were included in duplicate in all runs. To assess the repeatability of the new assay, two serum QC samples were measured in ten replicates through one run, and the intra-assay coefficient of variation (CV%) was calculated. Furthermore, the intermediate precision was determined by analyzing five replicates of two QC samples in two different runs.

The lower limit of quantification (LLOQ) and the limit of detection (LOD) were calculated based on a signal of 10 SD and 3 SD above the mean of 16 blanks, respectively. 89% of measured samples with the novel Ella assay exhibited GFAP values above the LLOQ. Parallelism was assessed in four endogenous samples (2 high and 2 low concentrations), diluted 1:2 to 1:8. Back-calculated concentrations were analyzed to determine the minimum required dilution (MRD). This approach aims to mitigate the matrix effects and ensure a reliable quantification of the endogenous GFAP. To test spike and recovery, two serum samples with low GFAP concentrations were diluted 1:2 (MRD defined in parallelism experiments) and divided into three aliquots. Subsequently, the samples were spiked with GFAP-free sample diluent, medium (100 pg/mL) and low (20 pg/mL) concentration of GFAP recombinant protein (Lyophilized Quality control, Simple Plex^TM^). The spiked volume was less than 10% of the final aliquot volume. Recovery was calculated in percentage. To test cross-reactions to highly abundant blood proteins, serial dilutions of two serum samples were spiked with physiological blood concentrations of human albumin (40 mg/ml) and immunoglobulin G (IgG) (10 mg/ml). Subsequently, GFAP levels were compared with unspiked samples. For the homemade Ella assay, 12 samples from the cohort measurements were excluded from the analysis as GFAP values were not measurable due to errors during measurements.

### GFAP measurement

The levels of serum GFAP in the clinical cohort were measured using the 2^nd^ gen GFAP blood assay developed by Biotechne on their microfluidic Ella platform (GFAP 2^nd^ gen assay, Biotechne, MN, USA). According to the manufacturer, the new Ella assay detects GFAP in a range of 2.52 - 9600 pg/mL. Serum samples were diluted using sample diluent SD13 (Biotechne, MN, USA) with a dilution factor of 1:2. Additionally, serum samples were also analyzed with the same microfluidic platform, using a homemade GFAP blood assay published by Fazeli et al. (20), with slight improvements. Finally, samples were measured with the Quanterix HD-X analyzer using the Simoa GFAP Discovery kit according to the manufacturer’s instructions (Qunaterix, MA, USA).

### Statistics

Data were analyzed and visualized using GraphPad Prism V.8.3.0 software (GraphPad Software, La Jolla, CA, United States). Mann-Whitney U-test and Kruskal-Wallis followed by Dunn’s post-hoc tests were applied to determine the significant differences between two or more groups. The Spearman correlation coefficients were calculated between GFAP levels obtained from different assays and it’s relation to age. The Bland - Altman method was carried out to assess the agreement between assays. A p-value < 0.05 was considered statistically significant.

## Results

### Performance of the 2^nd^ gen Ella assay

Validation experiments of the novel assay revealed an intra- and inter-assay CV of 10% and 12%, respectively. Dilution-adjusted concentrations of measured samples in the parallelism test were plotted (Fig. 1A), and a 1:2 dilution was chosen as the MRD. Considering the 1:2 dilution as an anchor, the further dilutions revealed a linear pattern. The relative error (%) of each sample was compared to the determined MRD of 1:2 with an accepted variation range of +/-25% in the following dilutions (Fig. 1B).

**Figure 1.**
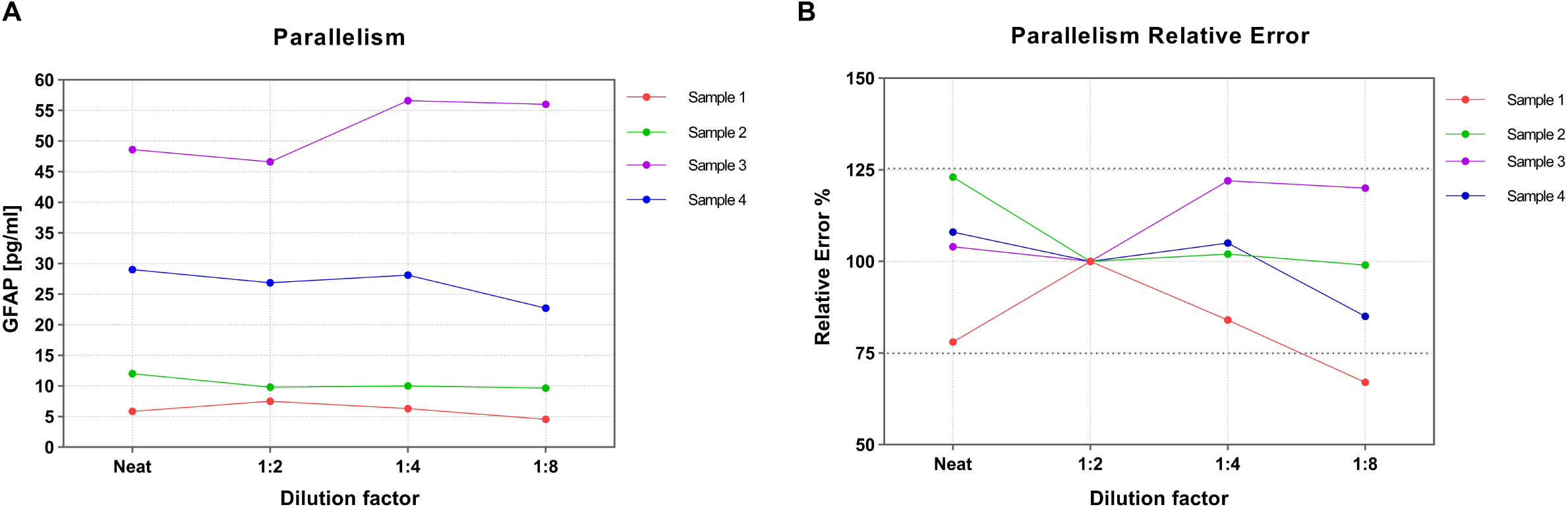
Parallelism assessments in four serum samples. (A) Back-calculated GFAP concentrations of four samples with low-, medium- and high GFAP concentrations. (B) Using 1:2 dilution as the MRD, the relative error in subsequent dilutions was within the accepted limitations of 75-125%. Sample 1 had a very low GFAP concentration and a dilution of 1:8 resulted in a near blank-level signal; therefore, a variation above 25% was observed. GFAP, glial fibrillary acidic protein; MRD, minimum required dilution.

The LLOQ and LOD were established at 2.8 pg/mL and 0.9 pg/mL, respectively. Stability assessments were carried out for both serum and CSF samples. The obtained data revealed that GFAP concentrations for both serum (Fig. 2A) and CSF samples (Fig. 2B) exhibited less than 20% variation when stored for up to 120 hours at either 4°C or room temperature (RT). Additionally, changes in serum GFAP concentrations remained within an acceptable range of ±20% after undergoing five freeze-thaw cycles (FTCs) (Fig. 2C), while the CSF GFAP levels exhibited a decrease following two FTCs (Fig. 2D). The spike and recovery experiment revealed a recovery of 82% and 85% for the low and high spike concentration, respectively. No evidence of a cross-reaction with human albumin or IgG was observed.

**Figure 2.**
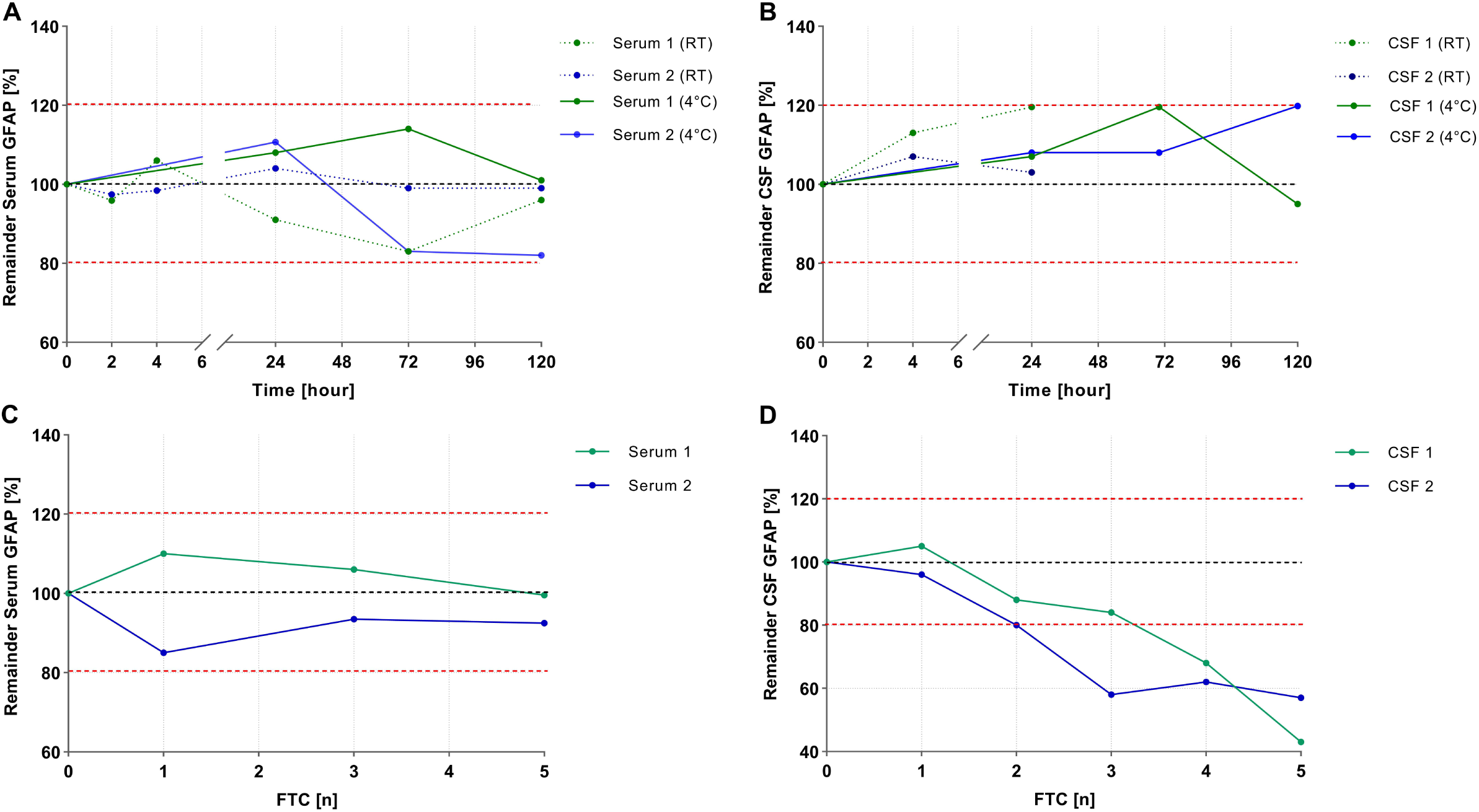
Evaluation of GFAP stability in serum and CSF. The stability of GFAP in serum and CSF was determined by comparing the relative content of GFAP in two serum samples (A) and two CSF samples (B) after storage at room temperature or 4°C, in comparison to the reference samples. Variations were found to be less than ±20%. Multiple freeze-thaw cycles were performed, and two serum (C) and CSF samples (D) were compared to the reference samples to assess GFAP’s relative concentration. Serum GFAP remained stable after undergoing up to five FTCs, while GFAP levels in CSF decreased after two cycles. CSF, cerebrospinal fluid, FTC, freeze-thaw cycle, GFAP, glial fibrillary acidic protein.

### Demographic and clinical features of the diagnostic groups

Implementing the 2^nd^ gen Ella GFAP blood, Simoa GFAP discovery, and microfluidic Ella homemade assay, 210 clinical serum samples from patients with AD (n=44), bvFTD (n=14), encephalitis (n=6), meningitis (n=9), meningoencephalitis (n=4), MS (n=38), and controls (n=95) were analyzed. According to the median, the control cohort was split into two groups: young (Y. Con, ≤50 years old) and old (O. Con, >50 years old). No significant differences existed in age between the O. Con and the AD, bvFTD, encephalitis, meningitis and meningoencephalitis group. Likewise, there was no significant age difference between the MS cohort and the Y. Con group. Table 1 provides an overview of the clinical and demographic characteristics of the diagnostic groups.

**Table 1.** Demographic and clinical parameters of the diagnostic groups.

|  | O. Con | AD | bvFTD | Enc | Men | ME | MS | Y. Con |
| --- | --- | --- | --- | --- | --- | --- | --- | --- |
| N | 48 | 44 | 14 | 6 | 9 | 4 | 38 | 47 |
| No. Female (%) | 24(50) | 28 (63) | 6 (43) | 2 (33) | 3 (33) | 2 (50) | 21 (55) | 26(55) |
| Age (year) | 61 (54-70) | 68 (64-75) | 64 (59-73) | 77 (59-79) | 50 (34-58) | 64 (51-79) | 32 (25-45) | 32 (28-43) |
| 2<sup>nd</sup> gen Ella Serum GFAP (pg/mL) | 6.78 (5.21-9.36) | 14.35 (10.98-19.41) | 7.85 (5.62-12.11) | 12.56 (8.04- 13.85) | 4.38 (2.92-5.77) | 6.17 (2.08-12.57) | 4.71 (3.24-7.46) | 3.32 (2.80-5.08) |
| Simoa Serum GFAP (pg/mL) | 155.1 (119.2-215.6) | 361 (249.2-496.3) | 225.2 (152.2-277.5) | 268.1 (169-333.9) | 122.7 (68.35-153.3) | 105.3 (54.75-226) | 115.2 (81.13-155.5) | 80.90 (57.80-111.9) |
| Homemade Ella Serum GFAP (pg/mL) | 5.52 (4.04-8.11) | 10.88 (8.38-13.04) | 6.01 (3.48-8.55) | 8.50 (5.52-13.42) | 4.56 (3.91-5.64) | 12.17 (4.09- 13.22) | 4.47 (3.49-6.01) | 4.12 (2.70-5.51) |

Age and concentrations are given as median with interquartile range in brackets. AD, Alzheimer's disease; bvFTD, behavioral variant frontotemporal dementia; Enc, Encephalitis; GFAP, Glial fibrillary acidic protein; Men, Meningitis; ME, Meningoencephalitis; MS, multiple sclerosis; O. Con, Old control; Simoa, Single-molecule array; Y. Con , Young control.

A similar pattern of positive correlation between age and GFAP concentrations, as determined with the three assays, was found in both the control and the entire cohort. The 2^nd^ gen Ella assay showed a correlation with age of r = 0.68 (95% CI: 0.59 – 0.74), p <0.0001) for the whole cohort and r = 0.60 (95% CI: 0.44 – 0.71), p <0.0001 for the controls only. All correlations with age can be found in the supplementary materials (Fig. S1)

### Cohort measurement

Comparing the GFAP assay results, the different diagnostic groups illustrated a similar GFAP concentration pattern (Fig. 3A–C). GFAP levels in AD patients were significantly higher than in the corresponding control group (O. Con) (p<0.0001 for all assays). In all three evaluations, the concentration of GFAP was considerably higher for AD patients compared to Meningitis patients (for 2^nd^ gen Ella and Simoa p<0.0001, homemade Ella p=0.0003).

**Figure 3.**
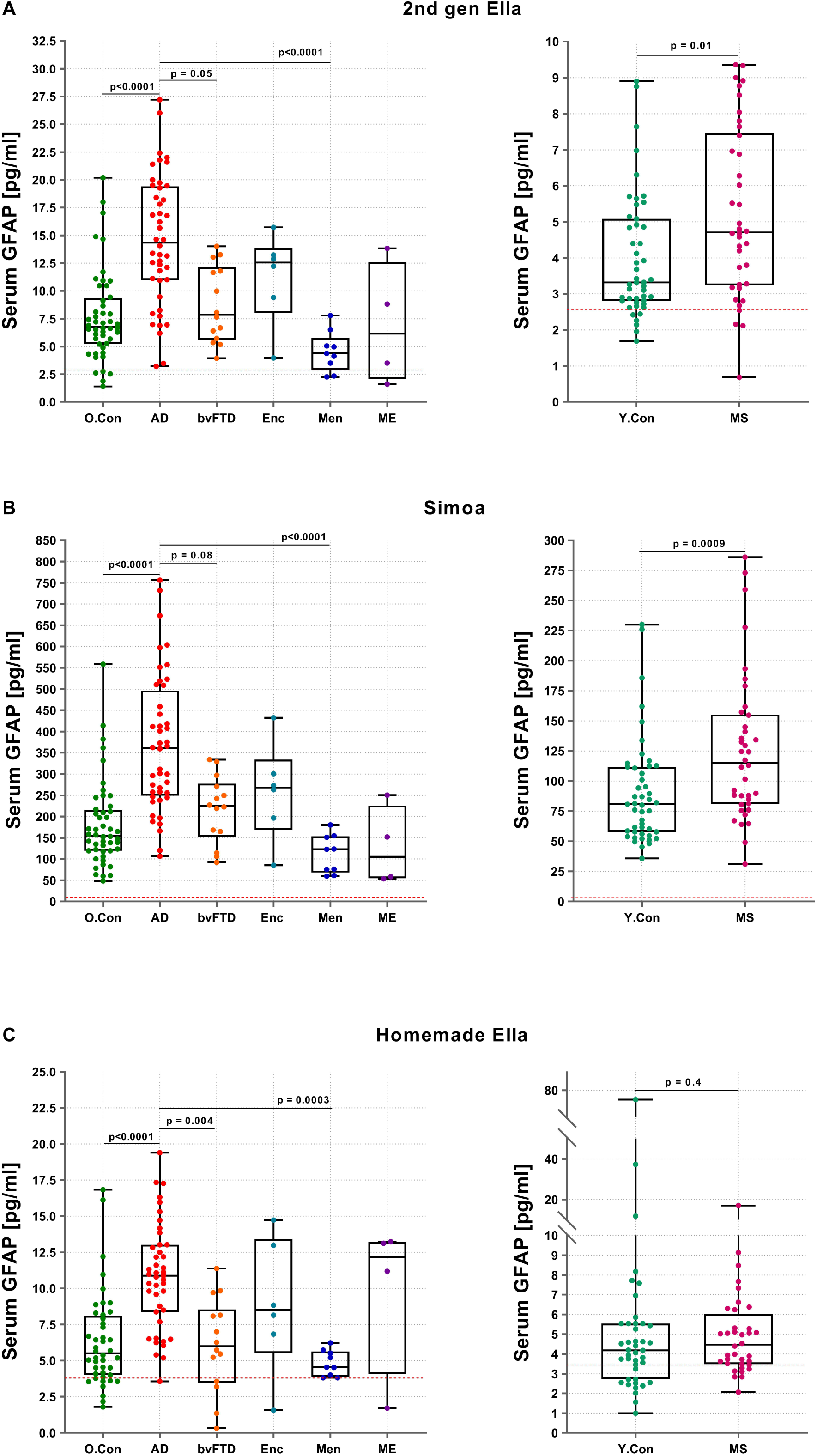
GFAP measurement in a clinical cohort using three different GFAP assays. (A) 2^nd^ gen Ella assay (n=210), (B) Simoa discovery kit (n=210), (C) homemade Ella assay (n=198). GFAP levels in the diagnostic groups were compared to the corresponding control cohort (Con) in the same age range. Boxes represent the median and interquartile range, with whiskers for minimum and maximum. The red-dotted lines represent the lower limit of quantification of each assay. The Mann-Whitney U-test was employed to compare GFAP levels between MS patients and young control patients. For the remaining comparisons, the Kruskal-Wallis test was initially conducted, followed by Dunn’s post-hoc test AD, Alzheimer’s disease; bvFTD, behavioral variant frontotemporal dementia; Enc, Encephalitis; GFAP, Glial fibrillary acidic protein; Men, Meningitis; ME, Meningoencephalitis; MS, multiple sclerosis; O. Con, Old control; Simoa, Single-molecule array; Y. Con, Young control.

Furthermore, measurements with the homemade Ella assay displayed significantly higher levels of GFAP in AD compared to bvFTD patients (p= 0.004). However, this difference was not significant for the other two assays (2^nd^ gen Ella p=0.05, Simoa p=0.08). In addition, the MS cohort in comparison to the associated control cohort (Y. Con), demonstrated significantly elevated GFAP levels in MS assessed by the 2^nd^ gen Ella assay (p=0.01) and Simoa (p=0.0009) but not with the homemade Ella (p=0.4). No significant differences in GFAP level were observed between the other diagnostic groups. For sensitivity comparison purposes, the LLOQ was added to the graphs and the number of samples below the LLOQ was calculated. For both the 2^nd^ gen and the homemade Ella assays, the percentage of samples with a concentration below the LLOQ was 11% and 23%, respectively. All samples analyzed with the Simoa were above the LLOQ.

### Method correlation

Serum GFAP levels in the whole cohort were highly correlated between the three assays (Fig. 4A-C). The strongest correlation was observed between the comparison of the novel 2^nd^ gen Ella and the Simoa assay (r=0.91, p<0.0001). Moreover, strong correlations were also observed between the two Ella assays (r=0.77, p<0.0001) and the Simoa-homemade Ella assay (r=0.74, p<0.0001). Simple linear regression of the 2^nd^ gen Ella and Simoa assay revealed an R^2^ of 0.86 and a slope of 24.

**Figure 4.**
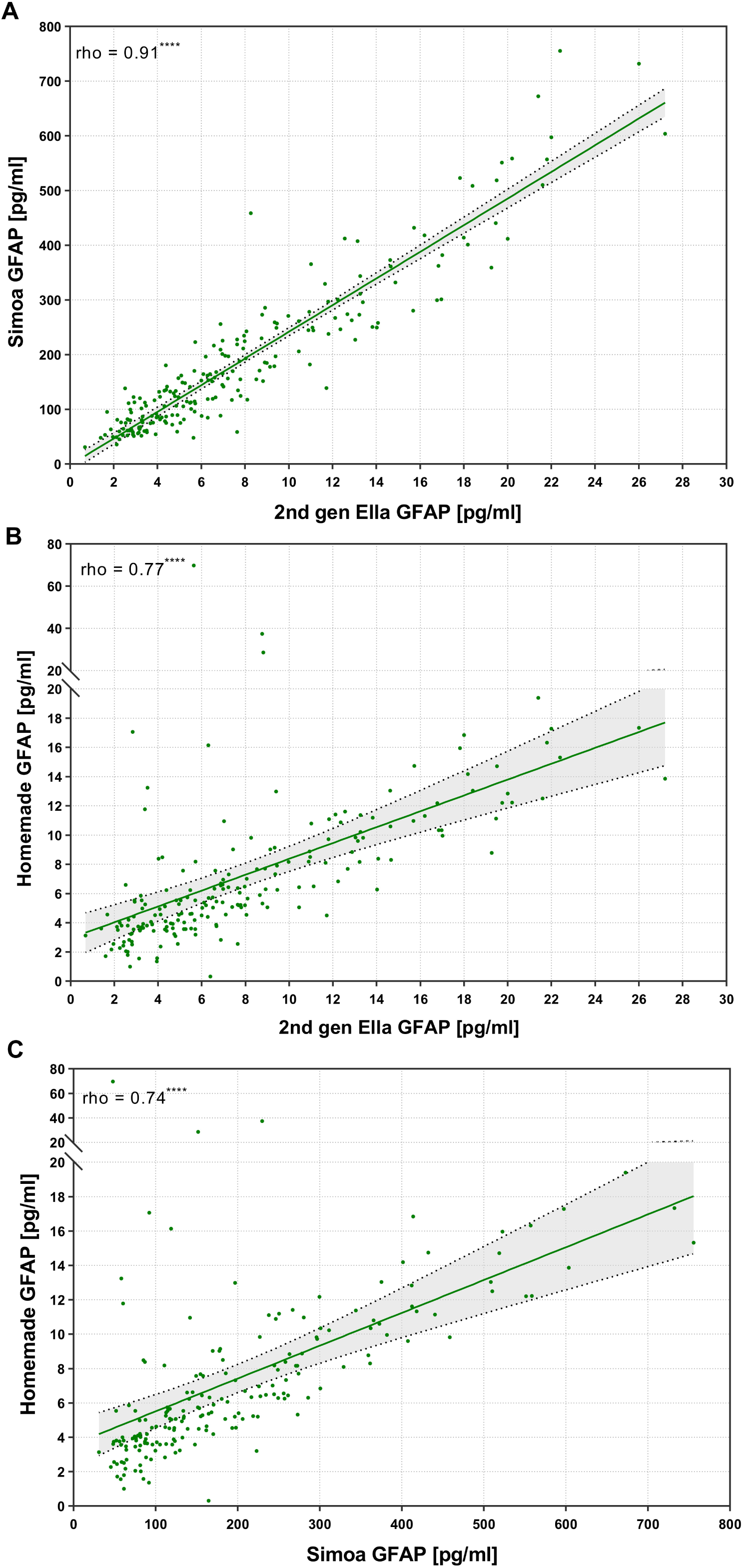
Pairwise correlations of serum GFAP measured by different assays. (A) Correlation between the novel 2^nd^ gen Ella assay and the Simoa assay in 210 samples with a correlation coefficient of r=0.91 (95% CI: 0.88 - 0.93, p<0.0001).(B) Correlation between the 2^nd^ gen Ella assay and the homemade Ella assay in 198 samples, with a correlation coefficient of r=0.77 (95% CI: 0.70 - 0.82, p<0.0001).(C) Correlation between the Simoa and the homemade Ella assay in 198 samples with a correlation coefficient of r=0.74 (95% CI: 0.67 - 0.80, p<0.0001). CI, Confidence interval; GFAP, Glial fibrillary acidic protein; Simoa, Single-molecule array.

Bland-Altman analysis depicted prominently higher absolute values for the Simoa compared to the Ella measurements (Fig. 5 & S 2A,B). Most of the observations were within the limit of agreement, as assessed by the confidence lines. Furthermore, the analysis revealed greater variability between assays in the lower detection range compared to the relatively more consistent results observed at higher GFAP levels.

**Figure 5.**
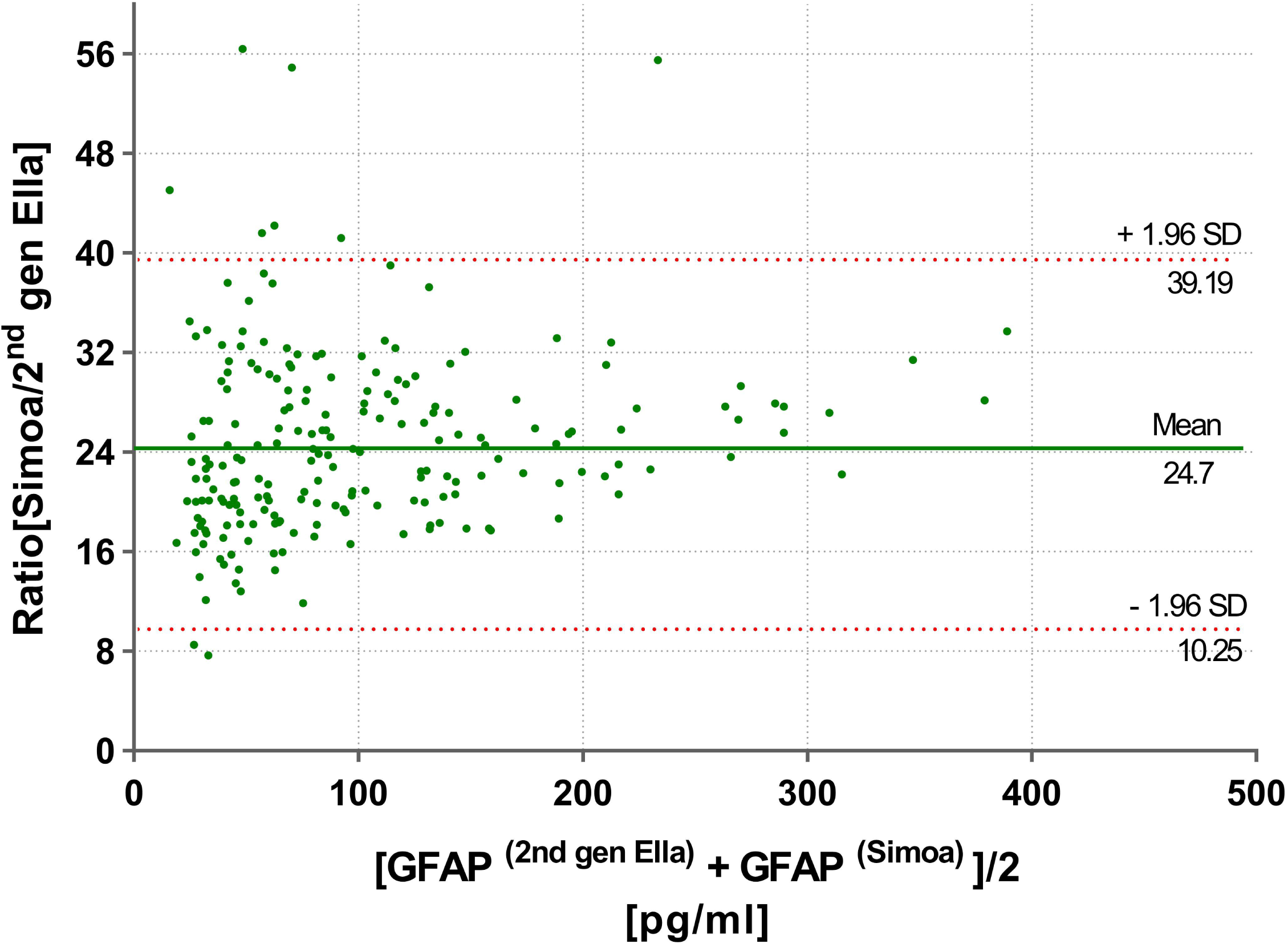
Bland-Altman analysis. Bland-Altman plot evaluates the agreement between the novel Ella and the Simoa assay. Circles illustrate the ratio of Simoa to 2^nd^ gen Ella GFAP for each sample (N = 210; Mean = 24.7 ; lower limit of agreement = 10.25; upper limit of agreement = 39.19). The 95% limits of agreement were displayed with horizontal red dotted lines in the graph, defined as the mean ratio of Simoa to 2^nd^ gen Ella GFAP values ±1.96 times the SD of the ratios. The solid green line represents the mean ratio. 2^nd^ gen Ella, second generation Ella; GFAP, Glial fibrillary acidic protein; SD, standard deviation; Simoa, Single-molecule array.

## Conclusion

In this study, we thoroughly validated the novel 2^nd^ gen GFAP Ella assay for its use in serum analysis. Additionally, we compared the results of a clinical cohort measurement with two existing blood GFAP assays and correlated the assays among each other. Validation experiments demonstrated a good precision for the 2^nd^ gen Ella assay, as both intra- and inter-assay CVs were clearly in an acceptable range below 15% (26). Moreover, the recovery rate of spiked GFAP protein was above 80%, suggesting a low interference of serum matrix effects.

The LLOQ and LOD of the 2^nd^ gen Ella assay were calculated to be 2.8 and 0.9 pg/mL, respectively. These values were higher than the Quanterix Simoa GFAP discovery kit (LLOQ: 1.3 pg/mL, LOD:0.2 pg/mL) and lower than the homemade Ella assay (LLOQ: 3.8 pg/mL, LOD 1.6 pg/mL, Fazeli et al., 2023, modified). In addition, all analyzed samples were above the LLOQ for the Simoa analysis (11% and 23% below the LLOQ for the 2^nd^ gen and homemade assay, respectively) demonstrating that while the 2^nd^ gen Ella GFAP offers greater sensitivity than the homemade assay its assay sensitivity is inferior to the Simoa. This might be due to the highly sensitive digital bead-based system used for Simoa analyses.

Furthermore, we also detected a huge difference between absolute GFAP concentrations, with markedly higher levels detected by the Simoa assay. This difference was further illustrated by the Bland-Altman plot, showing on average more than 20 times higher GFAP levels for Simoa. While we can only hypothesize the reason behind this, it is plausible that the discrepancy arises from the different antibodies used in each assay, potentially binding to different epitopes. Consequently, this could lead to the measurement of different GFAP isoforms or breakdown products, which could be present in the blood at different concentrations. A more straightforward explanation could be different calibrations of the standard curve. The latter might be more likely as we demonstrate a very strong correlation between the 2^nd^ gen Ella and Simoa, hinting at the measurement of the same GFAP proteins in the assays. Nonetheless, it is important to mention that the 2nd gen Ella and the Simoa assays cannot be used interchangeably.

The stability tests displayed stable serum GFAP concentrations after storage at either room temperature or 4°C for up to 120 hours and up to five freeze-thaw cycles. This robust stability facilitates sample handling in daily clinic use. As CSF GFAP concentrations are reported to decline after several FTCs using the Simoa and homemade GFAP assays (20,27), we decided to also include CSF in the stability tests of the 2nd gen GFAP Ella assay. CSF GFAP levels could sustain storage at 4°C and room temperature for up to 120 hours, but after two freeze-thaw cycles, they also began to decline using the 2^nd^ gen Ella assay. Therefore, we recommend using only fresh CSF samples for GFAP analysis when applying the 2^nd^ gen Ella assay.

Taken together, validation assessments demonstrated a good performance of the 2^nd^ gen Ella assay. For that reason, we proceeded to employ the assay for the analysis of a clinical cohort of 210 patients. Subsequently, we compared the results to two alternative GFAP assays. The obtained results revealed the same GFAP concentration pattern independent of the assay applied, suggesting the analysis of the same GFAP isoform or breakdown product. This is further strengthened by the strong correlation between the assays, especially the 2^nd^ gen and Simoa GFAP assay. The ratio Bland-Altman plot also demonstrated a good agreement among the assays. However, at lower GFAP concentrations, the assays exhibited increased variability, potentially due to the lower assays sensitivity in that concentration range.

Additionally, we assessed the correlation between GFAP levels and age within the control and the whole cohort, using the three assays. A strong and positive correlation was apparent in the results obtained from the 2^nd^ gen Ella and the Simoa assays when assessing both the control group and whole cohort which confirms previous studies (10,28).

When evaluating the differences in GFAP levels among the diagnostic groups, we also observed a prominent GFAP increase among individuals with AD compared to control patients, as evidenced by all three assays. These results align with previous research findings (7,8,10). For the comparison AD and bvFTD, literature reports a significant elevation in AD. In our study, we did only observe a clear trend to increased levels in AD but did not find a significant difference using the 2^nd^ gen Ella or Simoa. This is most likely due to the low number of bvFTD patients measured, and further studies using 2^nd^ gen Ella to investigate more samples need to be performed.

Comparison of GFAP levels between the MS cohort and the young controls demonstrated a significant increase in MS patient values measured by the 2^nd^ gen Ella and the Simoa assays, as has been shown in previous studies (29–32). However, no significant elevation was observed using the homemade Ella. A possible explanation may be the lower sensitivity of the homemade assay compared to the other two assays, leading to a higher overlap between MS and control patients. The observed trend to elevated levels in encephalitis patients proof the literature that GFAP is not a specific disease marker and inflammation of the brain parenchyma can already lead to increased blood GFAP levels (33).

The study’s limitations are primarily attributed to the small number of patients in some diagnostic groups, which restricts the ability to draw precise and definitive conclusions concerning their GFAP levels using the 2^nd^ gen Ella assay. This also limited the possibility of analyzing the different MS subgroups within the MS category. Despite these limitations, our study presents several notable strengths. First, we conducted a comprehensive validation of the novel microfluidic highly sensitive assay and confirmed its reproducibility, robustness, and reliability. Second, we measured GFAP levels in a well-characterized clinical cohort, including various neurological diseases. Third, we correlated the GFAP levels with two already established assays rendering it possible to compare the results with current GFAP literature.

In conclusion, our findings show the robustness and reliability of the novel 2^nd^ gen Ella assay for the quantification of serum GFAP. The assay displays a strong correlation with the currently most used GFAP blood immunoassays with the limitation of a lower sensitivity compared to bead-based approaches. Still the assay could be a cost-effective alternative for GFAP analysis and, due to its ease of use, has a strong potential to be applied in routine clinical GFAP measurements.

## Data Availability

All data produced in the present study are available upon reasonable request to the authors.

## Acknowledgments

We would like to thank all participating patients and the Bio-Techne Ella development team for providing the 2^nd^ gen Ella cartridges. Bio-Techne was not involved in any analysis or interpretation of data.

## Author contributions

Conception and design of the study: BF, SH, HT; sample collection and data management: BF, NGdSJ, PO, SJ, MS, DE, MO, SH, HT; study management and coordination: BF, SH, HT; statistical methods and analysis: BF, SH, HT; interpretation of results: BF, NGdSJ, PO, SJ, MS, DE, MO, SH, HT; manuscript writing (first draft): BF, SH, HT; All authors critically revised the manuscript and approved the final version.

## References

1. Yang Z, Wang KKW. Glial fibrillary acidic protein: From intermediate filament assembly and gliosis to neurobiomarker. Vol. 38, Trends in Neurosciences. Elsevier Ltd; 2015. p. 364–74.

2. Cabezas R, Ávila M, Gonzalez J, El-Bachá RS, Báez E, García-Segura LM, et al. Astrocytic modulation of blood brain barrier: Perspectives on Parkinson’s disease. Vol. 8, Frontiers in Cellular Neuroscience. Frontiers Research Foundation; 2014.

3. Middeldorp J, Hol EM. GFAP in health and disease. Vol. 93, Progress in Neurobiology. 2011. p. 421–43.

4. Brenner M. Role of GFAP in CNS injuries. Vol. 565, Neuroscience Letters. Elsevier Ireland Ltd; 2014. p. 7–13.

5. Pekny M, Pekna M. ASTROCYTE REACTIVITY AND REACTIVE ASTROGLIOSIS: COSTS AND BENEFITS. Physiol Rev [Internet]. 2014;94:1077–98. Available from: www.prv.org

6. Abdelhak A, Foschi M, Abu-Rumeileh S, Yue JK, D’Anna L, Huss A, et al. Blood GFAP as an emerging biomarker in brain and spinal cord disorders. Vol. 18, Nature Reviews Neurology. Nature Research; 2022. p. 158–72.

7. Benedet AL, Milà-Alomà M, Vrillon A, Ashton NJ, Pascoal TA, Lussier F, et al. Differences between Plasma and Cerebrospinal Fluid Glial Fibrillary Acidic Protein Levels across the Alzheimer Disease Continuum. JAMA Neurol. 2021 Dec 1;78(12):1471–83.

8. Oeckl P, Halbgebauer S, Anderl-Straub S, Steinacker P, Hussa AM, Neugebauer H, et al. Glial fibrillary acidic protein in serum is increased in Alzheimer’s disease and correlates with cognitive impairment. Journal of Alzheimer’s Disease. 2019;67(2):481–8.

9. Petzold A. Glial fibrillary acidic protein is a body fluid biomarker for glial pathology in human disease. Vol. 1600, Brain Research. Elsevier B.V.; 2015. p. 17–31.

10. Oeckl P, Anderl-Straub S, Von Arnim CAF, Baldeiras I, Diehl-Schmid J, Grimmer T, et al. Serum GFAP differentiates Alzheimer’s disease from frontotemporal dementia and predicts MCI-to-dementia conversion. J Neurol Neurosurg Psychiatry. 2022 Jun 1;93(6):659–67.

11. Bolsewig K, Hok-A-Hin YS, Sepe FN, Boonkamp L, Jacobs D, Bellomo G, et al. A Combination of Neurofilament Light, Glial Fibrillary Acidic Protein, and Neuronal Pentraxin-2 Discriminates Between Frontotemporal Dementia and Other Dementias. J Alzheimers Dis. 2022;90(1):363–80.

12. Kim KY, Shin KY, Chang KA. GFAP as a Potential Biomarker for Alzheimer’s Disease: A Systematic Review and Meta-Analysis. Vol. 12, Cells. MDPI; 2023.

13. Montoliu-Gaya L, Alcolea D, Ashton NJ, Pegueroles J, Levin J, Bosch B, et al. Plasma and cerebrospinal fluid glial fibrillary acidic protein levels in adults with Down syndrome: a longitudinal cohort study. EBioMedicine. 2023 Apr 1;90.

14. O’Connor A, Abel E, Benedet AL, Poole T, Ashton N, Weston PSJ, et al. Plasma GFAP in presymptomatic and symptomatic familial Alzheimer’s disease: a longitudinal cohort study. Journal of Neurology, Neurosurgery & Psychiatry [Internet]. 2023 Jan 1;94(1):90. Available from: http://jnnp.bmj.com/content/94/1/90.abstract

15. Sun M, Liu N, Xie Q, Li X, Sun J, Wang H, et al. A candidate biomarker of glial fibrillary acidic protein in CSF and blood in differentiating multiple sclerosis and its subtypes: A systematic review and meta-analysis. Mult Scler Relat Disord [Internet]. 2021 Jun 1;51. Available from: 10.1016/j.msard.2021.102870

16. Kim JS. Protein biomarkers in multiple sclerosis. encephalitis. 2023 Apr 10;3(2):54–63.

17. Hamilton CA, O’Brien J, Heslegrave A, Laban R, Donaghy P, Durcan R, et al. Plasma biomarkers of neurodegeneration in mild cognitive impairment with Lewy bodies. Psychol Med [Internet]. 2023 Jul 25;1–9. Available from: https://www.cambridge.org/core/product/identifier/S0033291723001952/type/journal_article

18. Bucci M, Bluma M, Savitcheva I, Ashton NJ, Chiotis K, Matton A, et al. Profiling of plasma biomarkers in the context of memory assessment in a tertiary memory clinic. Transl Psychiatry. 2023 Dec 1;13(1).

19. Gao F, Dai L, Wang Q, Liu C, Deng K, Cheng Z, et al. Blood-based biomarkers for Alzheimer’s disease: a multicenter-based cross-sectional and longitudinal study in China. Sci Bull (Beijing) [Internet]. 2023 Jul; Available from: https://linkinghub.elsevier.com/retrieve/pii/S2095927323004449

20. Fazeli B, Huss A, Gómez de San José N, Otto M, Tumani H, Halbgebauer S. Development of an ultrasensitive microfluidic assay for the analysis of Glial fibrillary acidic protein (GFAP) in blood. Front Mol Biosci. 2023;10.

21. Dubois B, Feldman HH, Jacova C, Hampel H, Molinuevo JL, Blennow K, et al. Advancing research diagnostic criteria for Alzheimer’s disease: the IWG-2 criteria. Lancet Neurol [Internet]. 2014 Jun 1 [cited 2023 Jan 19];13(6):614–29. Available from: https://www.thelancet.com/journals/laneur/article/PIIS1474-4422(14)70090-0/fulltext#.Y8kRInj9Hh4.mendeley

22. Thompson AJ, Banwell BL, Barkhof F, Carroll WM, Coetzee T, Comi G, et al. Diagnosis of multiple sclerosis: 2017 revisions of the McDonald criteria. Lancet Neurol [Internet]. 2018;17(2):162–73. Available from: http://europepmc.org/abstract/MED/29275977

23. Rabinovici GD, Miller BL. Frontotemporal lobar degeneration: Epidemiology, pathophysiology, diagnosis and management. Vol. 24, CNS Drugs. 2010. p. 375–98.

24. Rascovsky K, Hodges JR, Knopman D, Mendez MF, Kramer JH, Neuhaus J, et al. Sensitivity of revised diagnostic criteria for the behavioural variant of frontotemporal dementia. Brain. 2011;134(9):2456–77.

25. Venkatesan A, Tunkel AR, Bloch KC, Lauring AS, Sejvar J, Bitnun A, et al. Case definitions, diagnostic algorithms, and priorities in encephalitis: Consensus statement of the international encephalitis consortium. Clinical Infectious Diseases. 2013 Oct 15;57(8):1114–28.

26. Andreasson U, Perret-Liaudet A, van Waalwijk van Doorn LJC, Blennow K, Chiasserini D, Engelborghs S, et al. A practical guide to immunoassay method validation. Front Neurol. 2015;6(Aug).

27. Simrén J, Weninger H, Brum WS, Khalil S, Benedet AL, Blennow K, et al. Differences between blood and cerebrospinal fluid glial fibrillary Acidic protein levels: The effect of sample stability. Alzheimer’s and Dementia. 2022 Oct 1;18(10):1988–92.

28. Huebschmann NA, Luoto TM, Karr JE, Berghem K, Blennow K, Zetterberg H, et al. Comparing Glial Fibrillary Acidic Protein (GFAP) in Serum and Plasma Following Mild Traumatic Brain Injury in Older Adults. Front Neurol. 2020 Sep 18;11.

29. Abdelhak A, Huss A, Kassubek J, Tumani H, Otto M. Serum GFAP as a biomarker for disease severity in multiple sclerosis. Sci Rep. 2018 Dec 1;8(1).

30. Abdelhak A, Hottenrott T, Morenas-Rodríguez E, Suárez-Calvet M, Zettl UK, Haass C, et al. Glial Activation Markers in CSF and Serum From Patients With Primary Progressive Multiple Sclerosis: Potential of Serum GFAP as Disease Severity Marker? Front Neurol. 2019 Mar 26;10.

31. Schindler P, Grittner U, Oechtering J, Leppert D, Siebert N, Duchow AS, et al. Serum GFAP and NfL as disease severity and prognostic biomarkers in patients with aquaporin-4 antibody-positive neuromyelitis optica spectrum disorder. J Neuroinflammation. 2021 Dec 1;18(1).

32. Barro C, Healy BC, Liu Y, Saxena S, Paul A, Polgar-Turcsanyi M, et al. Serum GFAP and NfL Levels Differentiate Subsequent Progression and Disease Activity in Patients With Progressive Multiple Sclerosis. Neurology(R) neuroimmunology & neuroinflammation. 2023 Jan 1;10(1).

33. Saraste M, Bezukladova S, Matilainen M, Sucksdorff M, Kuhle J, Leppert D, et al. Increased serum glial fibrillary acidic protein associates with microstructural white matter damage in multiple sclerosis: GFAP and DTI. Mult Scler Relat Disord. 2021 May 1;50.

