## Supplemental File for "Quantification of blood glial fibrillary acidic protein using a second-generation microfluidic assay. Validation and comparative analysis with two established assays"

**Supplementary material**

**GFAP correlations with age**

The correlation between GFAP concentrations and age was assessed in both the control and whole cohort. Obtained results revealed a positive correlation in the control and the entire cohort with all three assays (Fig. S1 A-F). However, a stronger correlation was observed in entire cohort (2nd gen Ella : (spearman r = 0.68 (95% CI: 0.59 – 0.74), p <0.0001), Simoa: r = 0.70 (95% CI: 0.61 – 0.76), p <0.0001) compared to the control cohort (2nd gen Ella : r = 0.60 (95% CI: 0.44 – 0.71), p <0.0001, Simoa: r = 0.65 (95% CI: 0.50 – 0.75), p <0.0001). Although this was less clear for the homemade Ella assay in the total cohort (r = 0.56 (95% CI: 0.45 – 0.65), p <0.0001) (Fig. S1 E) and in the control cohort (r = 0.35 (95% CI: 0.14 – 0.52), p =0.0009) (Fig. S1 F), in comparison with the other two assays.

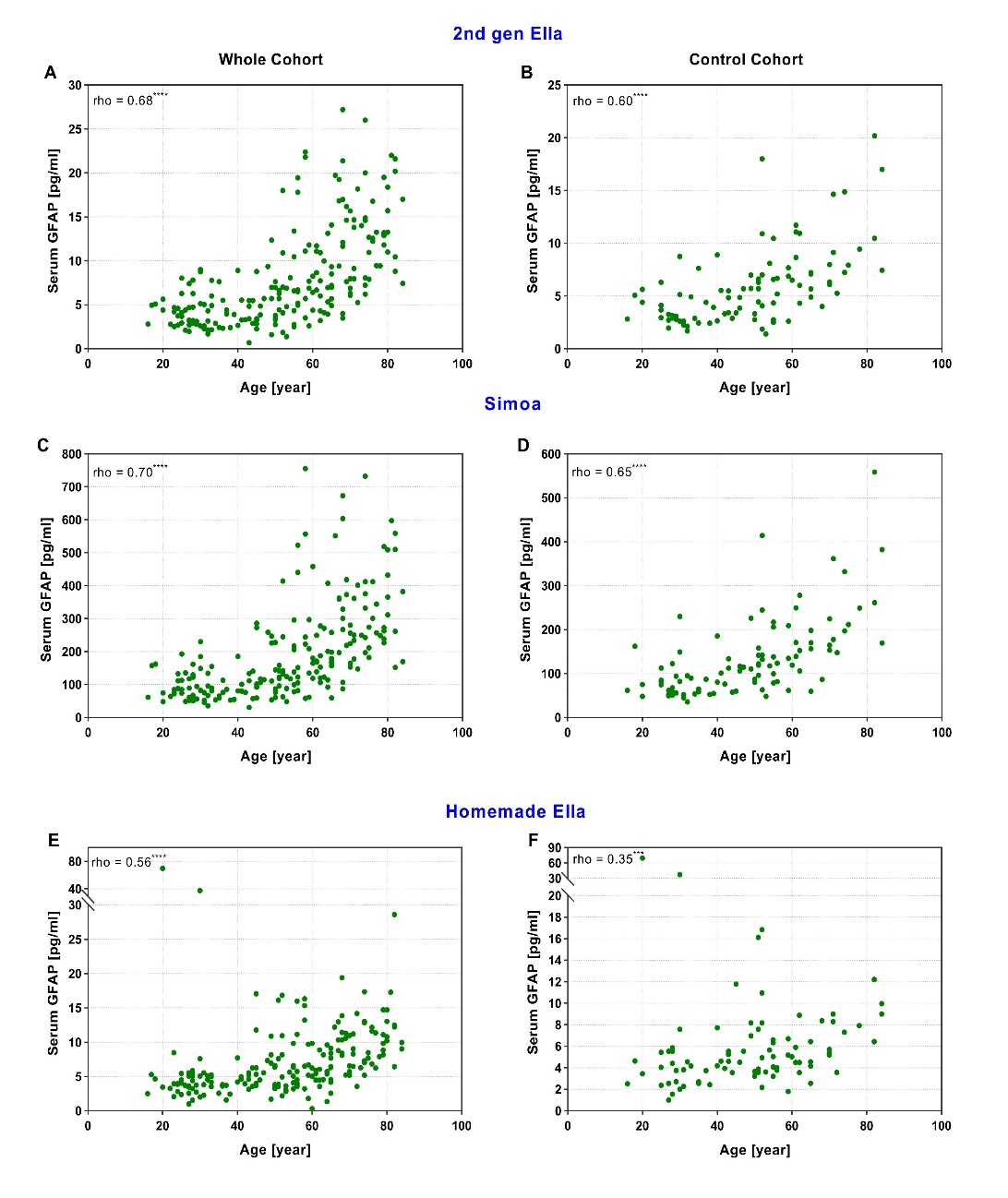

***Figure S1. Assessment of correlation between GFAP concentration and age.***

*Spearman correlation (rho) between age and serum GFAP measured using the 2^nd^ gen Ella assay in the whole cohort* *(n=210) (A) and the control cohort (n=95) (B), the Simoa assay in the whole cohort (n=210) (C) and the control cohort (n=95) (D), and the homemade Ella assay in the entire cohort (n=198) (E) and the control cohort (n=86) (F). GFAP, Glial fibrillary acidic protein; Simoa, Single-molecule array.*

**Correlation and agreement between assays**

Bland Altman plots confirmed agreement between measured GFAP concentrations in a paired comparison of 2^nd^ gen Ella -homemade Ella (Fig. S 2A) and Simoa - homemade Ella (Fig. S 2B) assays. Only one measurement was outside the limit of agreement as assessed by the 95% confidence lines.

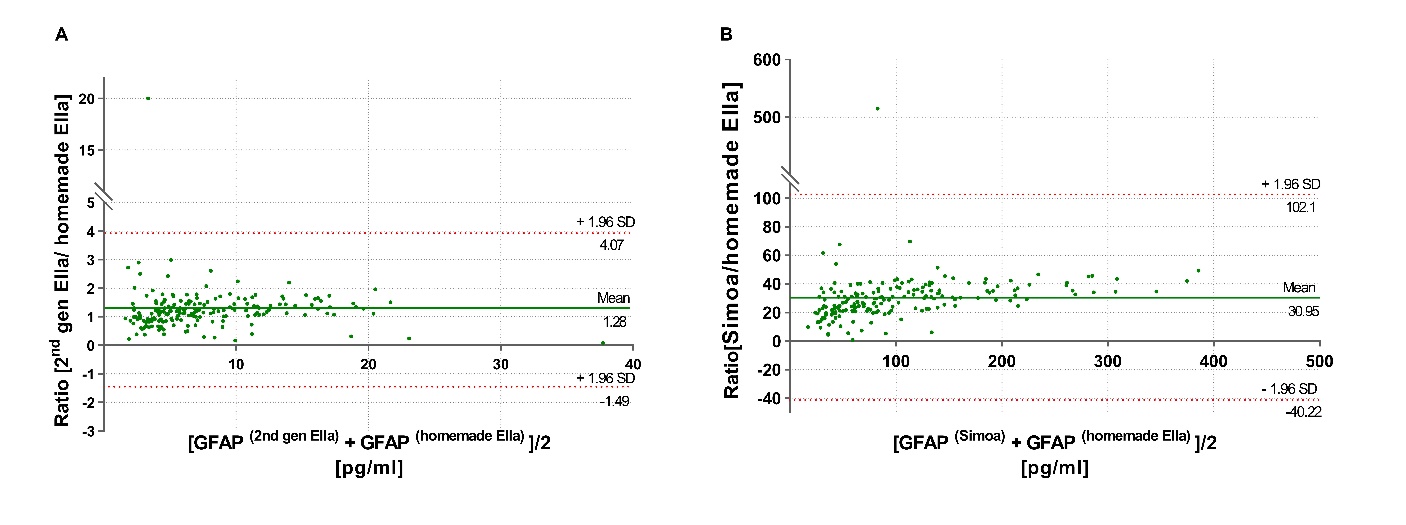

***Figure S 2. Bland Altman analysis***

*(A) Bland-Altman plot comparing agreement between GFAP concentrations determined using 2nd gen and homemade Ella assays. (B) Bland-Altman plot comparing agreement between GFAP concentrations determined using Simoa and homemade Ella assays. The solid green line represents the mean ratio between two assays and the dashed red lines display 95% limits of agreement. 2^nd^ gen Ella, second generation Ella; GFAP, Glial fibrillary acidic protein; SD, standard deviation; Simoa, Single-molecule array.*
